# Continuous Patient State Attention Models

**DOI:** 10.1101/2022.12.23.22283908

**Authors:** Vinod K. Chauhan, Anshul Thakur, Odhran O’Donoghue, Omid Rohanian, David A. Clifton

## Abstract

Irregular time-series (ITS) are prevalent in the electronic health records (EHR) as the data is recorded in EHR system as per the clinical guidelines/requirements but not for research and also depends on the patient health status. ITS present challenges in training of machine learning algorithms, which are mostly built on assumption of coherent fixed dimensional feature space. In this paper, we propose a computationally efficient variant of the transformer based on the idea of cross-attention, called Perceiver, for time-series in healthcare. We further develop continuous patient state attention models, using the Perceiver and the transformer to deal with ITS in EHR. The continuous patient state models utilise neural ordinary differential equations to learn the patient health dynamics, i.e., patient health trajectory from the observed irregular time-steps, which enables them to sample any number of time-steps at any time. The performance of the proposed models is evaluated on in-hospital-mortality prediction task on Physionet-2012 challenge and MIMIC-III datasets. The Perceiver model significantly outperforms the baselines and reduces the computational complexity, as compared with the transformer model, without significant loss of performance. The carefully designed experiments to study irregularity in healthcare also show that the continuous patient state models outperform the baselines. The code is publicly released and verified at https://codeocean.com/capsule/4587224.

## I. Background

Uneven time intervals between measurements of a patient’s attributes, such as heart rate, lead to irregularity in the electronic health records (EHR), which results in missing values while preparing the data for processing with machine learning models [2]. In EHR, irregular time-series (ITS) occur due to several reasons. For example, the data in EHR system is not recorded for the research purposes but is recorded as per the guidelines, medical requirements and for supporting medical claims etc., and all the measurements and treatments are dependent on the patient health status [3]. ITS are widely prevalent in primary and secondary care, including critical care, e.g., the MIMIC-III dataset has a missing rate of over 90% for hourly sampled ITS [4].

The adoption of EHRs in healthcare has presented great opportunities to develop machine learning (ML) models and artificial intelligence (AI) techniques to reduce the workload on an already burdened healthcare system, guide clinical decision making and increase efficiency of healthcare resources [5], [6]. However, ML models are, mainly, based on assumption of coherent fixed-dimensional feature space and the presence of irregularity in EHRs invalidates that assumption. The irregularity presents challenges to train the ML models without affecting performance on downstream tasks [7]. As a result, it is crucial to develop techniques for the correct imputation of missing time-steps in EHR for a number of reasons, including resource management, triaging, diagnosis, treatment, and prognosis.

Due to the importance and wide prevalence of ITS in health-care, ITS and the resulting missing values in EHR have received increasing attention from the research community, and there has been extensive research to address the irregularity, e.g., [2], [3], [8]–[10]. A wide variety of techniques have been proposed to handle ITS, e.g., from the traditional statistical techniques for replacement of missing values (such as using mean and median values etc.), imputation, interpolation, and matrix completion-based techniques [11] to advanced methods, such as neural processes [3], modification of recurrent neural networks [12], neural ordinary differential equations (NODE) [8], and attention based techniques [9] etc.

Traditional basic statistical techniques for replacing missing values, such as zero, mean, median, and carry forward, are biased and make strong assumptions about the underlying data generation process. This is reported to result in loss of performance in downstream prediction tasks [13]. Many other modern techniques for modelling fail to capture feature-correlations in time-series [12], separate the modelling of missingness from the downstream task, and fail to learn the missingness pattern, or are not adequately efficient to handle long sequences, noise and multi-modality of the data [11] etc. Moreover, there are scarce techniques that can handle completely missing time-steps, such as in [8] and others mostly handle partially missing values, such as in [2].

In this paper, our contributions are two-fold: first we propose a computationally efficient variant of the transformer [15] based on the idea of cross-attention [16], [16], [17], called Perceiver, to process long time-series in EHR, and second, we propose a continuous variant of these attention based models, i.e., the Perceiver and the transformer to deal with ITS and address the above limitations of ITS techniques. The proposed continuous patient state attention models learn the patient health trajectory for end-to-end learning from the ITS in EHRs, which can handle long sequences, noise and completely missing time-steps, including the sparse time-series.

The transformer based models are one of the most successful deep learning techniques, which have shown great results across different domains [2], [18]. However, the quadratic dependence of the transformer based models on the input limits their application to long sequences. To address this issue, recently, Perceiver based models [16], [16], [17] proposed the idea of cross-attention to squeeze the large inputs to tighter learnable latents, which are then followed by self-attention operations (the transformer) on the squeezed inputs. We also utilise the idea of cross-attention to propose Perceiver model for EHR to handle the long sequences of time-series, as discussed in the Section II. The Perceiver could be very valuable in healthcare since EHRs data represent a lot of information about the patients, and working with the complete and long trajectory of patient health status can yield very good results.

To address the irregularity in EHR data, we propose continuous variants of the Perceiver and the transformer for the patient health status, called as COntinuous Patient state PERceiver (COPER) and Continuous Transformer (CTransformer), respectively. These continuous state models learn the patient health dynamics, i.e., patient trajectory from the observed irregular time-steps from which any point can be sampled and used to generate a regular time-series to be processed with the Perceiver/transformer model. COPER/CTransformer can handle the completely missing time-steps, i.e., time-steps where no data is recorded, as well as small noise in the observations because it can generate the complete time-series after learning from the observed irregular time-steps. COPER/CTransformer achieve continuity in the patient health status using embedding and NODE. The continuous patient states could be helpful in a wide range of tasks including diagnosis, prognosis and treatment and in disease progression modelling.

The proposed work have some similarities to [8], [14] and [16], [17], [19]. The work in [8], [14], specifically [8] proposed latent ODE (LODE) based on recurrent neural networks and develops an encoder and decoder based architecture employing NODE in both to address the irregularity. LODE learns the dynamics of hidden state. Thus, our work is different from the LODE, in terms of using non-recurrent neural networks, using one NODE and using NODE for continuity of patient state rather than hidden state of the neural network. Moreover, the works in [16], [17], [19] are based on the idea of cross-attention of inputs with learnable latents for reducing the complexity of the transformer based architectures. Our work also borrows the idea of cross-attention but is architecturally different (refer to Section II for details) from the existing work, and also used for solving a different problem.

To evaluate the empirical performance of the proposed techniques, we have used in-hospital-mortality (IHM) prediction task using MIMIC-III and Physionet-2012 datasets, which contain time-series data from the intensive care unit (ICU), and using area under the receiver operating curve (AUROC) and area under precision recall curve (AUPRC) as the performance metrics. The Perceiver is compared with the long short-term memory (LSTM) and temporal convolutional network (TCN) as baselines. For evaluating the performance of the continuous patient state models, we have designed experiments to study the irregularity at 25%, 50% and 75% missing time-steps by randomly removing the time-steps, and compared them with simple baselines, like LSTM and Perceiver with carry forward, and advanced state-of-the-art techniques, like LODE [8] and Multi-Time Attention Network (mTAND) [9].

The contributions of the paper are summarised below.

- A computationally efficient variant of the transformer, called Perceiver, is proposed for time-series in EHR data, which is based on the idea of cross-attention of the inputs with *tighter* learnable latents. The cross-attention operation helps to reduce the computations by squeezing the long sequences to smaller latents. Perceiver presents another potential alternative for processing time-series in EHR and enables the processing of long patient trajectories.
- To address the irregularity in EHR data, continuous variants of the Perceiver and the transformer, called COPER and CTransformer, respectively, are proposed, which learn the dynamics of the patient health status from the irregular observed time-steps using neural ordinary differential equations. The proposed continuous patient state attention models can handle long sequences, noise and completely missing time-steps, including the sparse time-series. The continuous patient states could be helpful in a wide range of tasks including diagnosis, prognosis and treatment and in disease progression modelling.
- Empirical evaluation of the proposed techniques is performed on in-hospital-mortality prediction task using MIMIC-III and Physionet-2012 datasets with AUROC and AUPRC as performance metrics. The experiments show that the Perceiver can be used as a potential alternative for processing time-series in EHR, and significantly reduces the computations as compared to the transformer without significant loss of performance. The specifically designed experiments for continuous patient state models also show their efficacy to deal with the irregular time-series in EHR. Moreover, the proposed techniques also employ predictive uncertainty to improve transparency and trustworthiness, and is used to communicate uncertain cases to the experts to avoid making uncertain decisions.

The preliminary idea and the results of the present work were published in a four page paper [1]. This paper revises the idea and the empirical evaluation of [1] in a number of ways: (i) methodology is discussed in details, with the addition of algorithmic details, (ii) the architecture/idea is revised to have continuity only in the patient health state as this is sufficient to address the irregularity in EHR, which also improves the results than having continuity in the input as well as output spaces. Moreover, in the style of the continuous Perceiver, continuous version of the transformer is also proposed, (iii) new experiments are added to show the utility of the Perceiver over the transformer architecture, (iv) additional dataset and metric are considered to evaluate the proposed techniques, and (v) experiments are added to utilise the uncertainty of the proposed techniques to improve transparency and hence trustworthiness.

## II. Methods

In this section, we discuss the architecture and algorithmic details of the proposed Perceiver and the continuous patient state models.

### A. Perceiver

Transformer [15] based models have been successful across different domains with different modalities, including time-series in healthcare [20]. However, the main limitation of these models is their quadratic dependence on the input size, which results in large computational complexity when dealing with long context inputs, limiting their applicability to such problems that are quite common in healthcare time-series data [21].

Perceiver [16] based models are recent advancements to the transformer [15] based models and they address the issue of quadratic dependence of the transformers on the input by introducing cross-attention operation of learnable smaller latents with inputs. The cross-attention distils the long sequence input to smaller latents which is followed by self-attentions (transformer) on the squeezed latents, as given below. In our time-series settings, a long sequence of time-steps can be squeezed into a customised number of latents for processing with the transformer based models, which otherwise could be computationally very expensive or even infeasible in some cases to use the transformers directly on the input data.

The architecture of the COPER model, and the Perceiver as a component of the COPER, is presented in Fig. 1. The proposed Perceiver model borrows the idea of cross-attention from the original Perceiver based models [16], [17], [19] but has different architecture as shown in the Fig. 1. The Perceiver uses cross-attention operation to squeeze the input sequence length from *T* time-steps into *l < T* number of latents of same feature dimension as original sequence. The cross-attention is applied *M* -times on the input and the outputs are averaged, which then can be processed using transformer (self-attention) layers leading to lower computations as compared with processing the original input directly with the transformers.

**Fig. 1:**
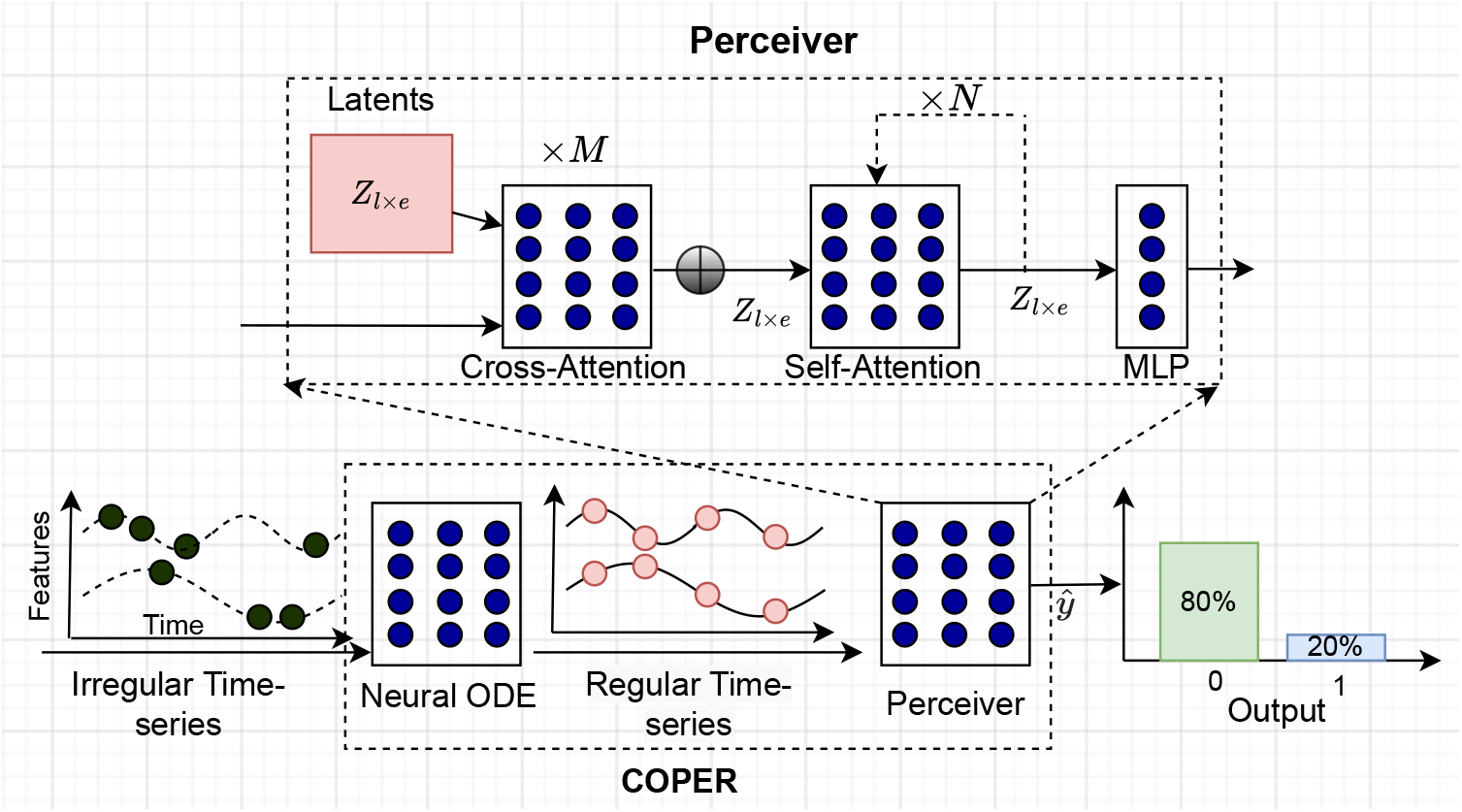
Architecture of COPER: An embedding of the irregular time-series is passed through the NODE, which captures the patient dynamics from the observed time-steps, and is used to generate a regular time-series. The generated regular time-series is then fed to the Perceiver model which first squeezes the long sequence of *T* time-steps to a *l < T* latents using the cross-attention and then followed by self-attention operations.

Suppose 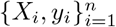 be the training dataset with *n* patients where 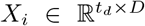 represents ITS of a patient *i* having *D* features for uneven *t*_*d*_ time-steps recorded for feature *d*, where each time-step represents the health status of the patient. *y*_*i*_ ∈ {0, 1} represents the patient outcomes (say in-hospital-mortality where 0 refers to patient lives to be discharged otherwise dies in the hospital). First, let’s define the attention operation [15], which is a scaled dot product attention between a set of queries (Q), keys (K) and values (V), as given below.

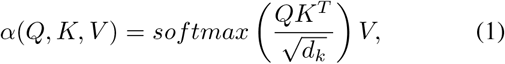

where *α* denotes the attention function and *d*_*k*_ is the dimension of the key vector. The self-attention operation has *Q* = *K* = *V* = *X*_*i*_ while cross-attention operation has *Q* = *Z* and *K* = *V* = *X*_*i*_, where *X*_*i*_ *∈ ℝ*^*T* ×*e*^ represents a data point (a patient in our case) with *e* features having *T* time-steps, and *Z* ∈ *ℝ*^*l*×*e*^ for 1 ≤ *l* ≤ *T* number of latents. Algorithm 1 provides details about the flow of information through the COPER and the Perceiver, which is explained in the next subsection.

#### Algorithm 1 COntinuous Patient state PERceiver

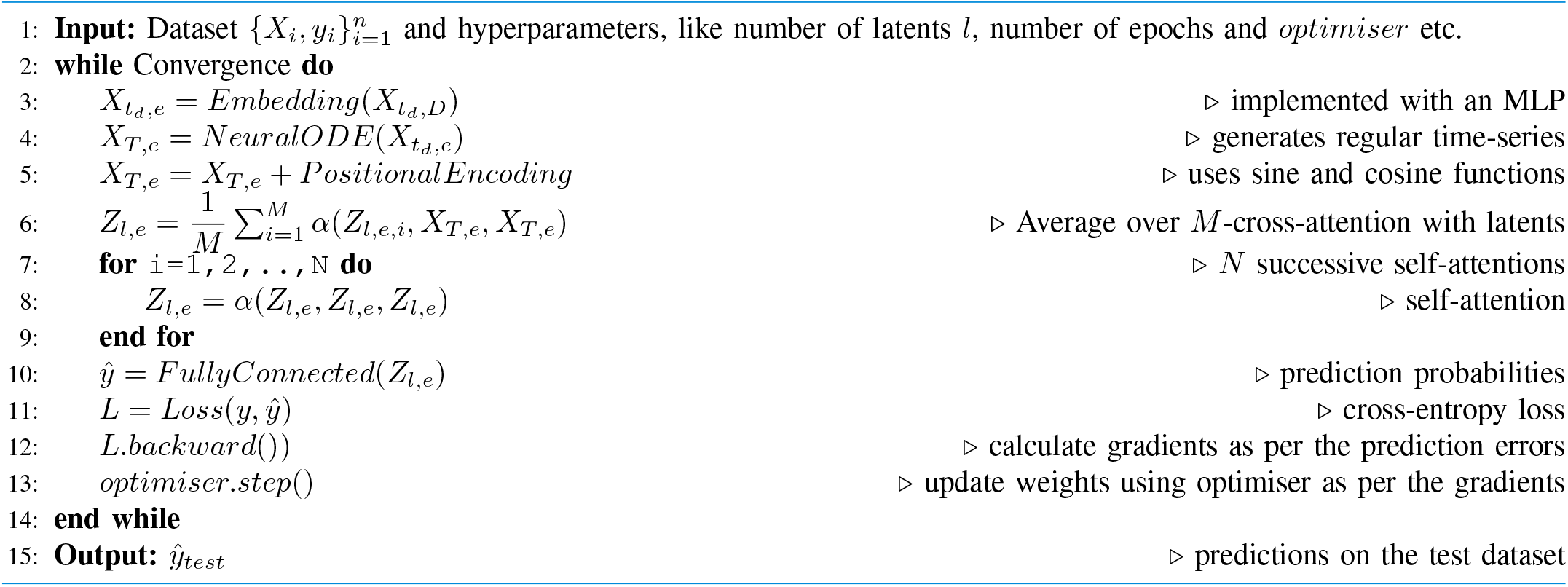

### B. Continuous Patient State Attention

Continuous patient state attention models are an advanced deep learning models to handle irregular time-series data in EHRs. They learn the patient health trajectory from the observed time-steps, i.e., observations of the patient health status at uneven time-steps. By learning the patient health dynamics, they can handle the irregularity as well as noise to some extent in the patient health status for successfully predicting the patient health outcomes.

COPER is based on the recent advancements of neural ordinary differential equations (NODE) and the Perceiver models to handle the ITS in EHRs, and can be applied to different tasks. The overall architecture of the COPER model is represented in Fig. 1 and the pseudocode for the representing the flow of information is given in the Algorithm 1.

As described in the algorithm, COPER processes the input ITS 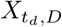, having *D* features with *t*_*d*_ time-steps for feature *d*, by first learning an optional embedding 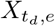 of size *e* for each time-step, using a single layer multilayer perceptron (MLP). These embeddings are then processed with the NODE [14], which are another recently developed class of neural networks. NODE helps to capture the dynamics of patient health status from which patient health status can be inferred at any time and a regular time-series can be generated. NODE consists of a neural network and a black-box ordinary differential equation (ODE) solver. The neural network outputs derivative of the patient health status, which is fed to an ODE solver. The ODE solver enables the model to calculate the patient health status at any time step, resulting in a continuous space, as described below.

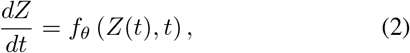

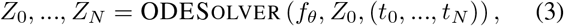

where *Z* is a patient state, *f*_*θ*_ is a neural network which parameterises the derivative of patient state. The ODESolver takes the derivative from the *f*_*θ*_ and initial patient state *Z*_0_ and calculates the patient state at the desired time-steps (*t*_0_, …, *t*_*N*_).

Steps 5-10 are part of the Perceiver model, which first adds positional encoding to the input for maintaining the order information of the time-steps using sine and cosine functions, as given below [15]:

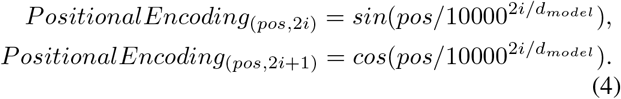

As shown in the Fig 1, the Perceiver applies *M* cross-attentions on the encoded input with the latents *Z*, followed by *N* successive self-attention operations on the average of cross-attention operations (*M* = 1, *N* = 1 in our experiments). The resulting output is then passed through a fully connected layer to predict the output probabilities, and followed by a standard machine learning process to update the parameters.

An architecture and algorithm for CTransformer can be obtained by replacing the cross-attention operation with the self-attention in the architecture and algorithm of COPER.

## III. Results

This section presents details about prediction task, datasets, performance metrics, baselines and experiments.

### A. Datasets and Baselines

The proposed models are evaluated for in-hospital mortality (IHM) prediction task using two publicly available datasets, i.e., Physionet Challenge 2012 dataset (hereon referred to as Physionet) [22], [23] and Medical Information Mart for Intensive Care (MIMIC-III) dataset (hereon referred to as MIMIC) [24]. These are time-series dataset based in the intensive care unit (ICU) setting. IHM is a binary classification task to predict from the first 48 hours of ICU admission for hourly data if patient will die in the hospital or live to be discharged. IHM prediction is very important for resource management, triage, initial risk assessment and designing effective treatment plans [12]. For preprocessing of MIMIC dataset, we have followed [25] to get a dataset with 76 features and 14,681, 3,236 and 3,222 samples in train, validation and test datasets^1^, respectively. For Physionet dataset, we follow the preprocessing as used in [8]. The dataset has 47 features and a total 8,000 samples. Due to smaller size, we have used 5-fold cross-validation in our experiments. Furthermore, the validation data is taken as 20% of the training data.

Two sets of experiments are designed as follows: first set presents the Perceiver model – a computationally efficient variant of the transformer – as a potential alternative for learning from time-series data. The experiments compared the model with Long Short-Term Memory (LSTM) [26] and Temporal Convolutional Network (TCN) [27] – the widely used techniques for handling time-series data. Experiments are also designed to show how the latents in Perceiver can be used to squeeze the long sequences into tight smaller number of latents to reduce the computational cost. Second set of experiments present continuous patient state attention models for handling the irregularity in EHRs. Since our proposed work is based on attention and NODE so for comparative study, we have chosen simple baselines as well as baselines based on state-of-the-art attention and NODE based techniques. The selected baselines are LSTM and Perceiver with carry forward to deal with the missing steps, and Multi-Time Attention Network (mTAND) [9], and latent ODE (LODE) [8] which are advanced state-of-the-art techniques for handling irregularity and are based on attention and NODE, respectively. To study the irregularity, we have designed experiments at 0%, 25%, 50% and 75% irregularity by randomly removing the time steps. Area under the receiver operating curve (AUROC) and area under the precision recall curve (AUPRC) are used as performance metric for the comparative study.

### B. Experimental Settings

Parameters of COPER are selected using a random search, and trial and error over a range of values: embedding layer is implemented using a multi-layer perceptron (MLP) with single hidden layer of 32 (16, 32, 64, 128) neurons (where values inside parenthesis represent the set of values tried), NODE are implemented using an MLP with three hidden layers of 128 (50, 100, 128) neurons for each NODE, cross- and self-attention heads have 128 (32, 64, 128, 256) dimensions, latents have 64 (32, 64, 128, 256) dimensions, dropout for attentions and NODE networks are set to 0.5 (0.2, 0.3, 0.4, 0.5, 0.6). The number of latents, unless specified, are set equal to number of time-steps. Number of cross-attention operation is set to one, i.e., *M* = 1 for simplicity, and number of self-attention operations are *N* = 1(1, 2, 3, 4, 5). For LSTM, number of layers are set to two (one, two) each with hidden state of size 50 (16, 32, 50, 64, 128), dropout rate is set to 0.5 (0.2, 0.3, 0.4, 0.5, 0.6) and single-directional (single, bi). The TCN implementation and hyperparameter setting is followed from [27] with a dropout rate of 0.70%^2^. For mTAND^3^, we follow the source paper and have set the hyperparameters as (Physionet, MIMIC): alpha (100, 5), learning rate (0.0001, 0.0001), rec-hidden dimension (256, 256), gen-hidden dimension (50, 50), latent-dimension (20, 128), norm (true, true), kl (true, true), learn-emb (true, true), k-iwae (1, 1), and number of epochs are set to 300. For the LODE^4^, we follow the source paper and have set the hyperparameters as (Physionet, MIMIC): latent-dimension (20, 40), rec-dimension (40, 80), poisson (true, true), and number of epochs are set to 300.

For all the models, we have set the optimiser to Adam [28] with a constant learning rate of 0.0001. To avoid overfitting, in addition to dropout, we have used early stopping with a patience of 10 epochs. Total number of epochs are set to 100 unless provided by the baseline paper. The batch size is set to 64 for all the models except LODE where it is set to 32 with MIMIC dataset because LODE is memory intensive and the machine crashes with 64 data points. Each experiment is executed with five seed values. All the experiments are implemented in Pytorch [29] and executed on an Ubuntu machine (64GB RAM, 1 NVIDIA GeForce GPU 12 GB). The code is publicly released and verified at https://codeocean.com/capsule/4587224.

### C. Perceiver

We compared the Perceiver model against LSTM and TCN – the widely used techniques for the time-series data. Fig. 2 presents the comparative study on Physionet dataset using the AUPRC and AUROC as the performance metrics. Upper panel presents results using the AUPRC, and we observe that Perceiver significantly outperforms the LSTM (*p <* 0.001). TCN performs the worst in terms of AUPRC on Physionet. The outlier performance in all the models are present due to cross-validation, as the performance of the models is better on one of the folds compared to the rest. Lower panel of the figure, presents results for the AUROC metric, and we observe results similar to the AUPRC. We find that Perceiver significantly outperforms the LSTM (*p <* 0.001) and TCN, once again, performs the worst. The variance in the values of the Perceiver model is the least among all the models. Thus, the Perceiver model exhibits significantly better performance on the Physionet dataset for the in-hospital-mortality prediction task.

**Fig. 2:**
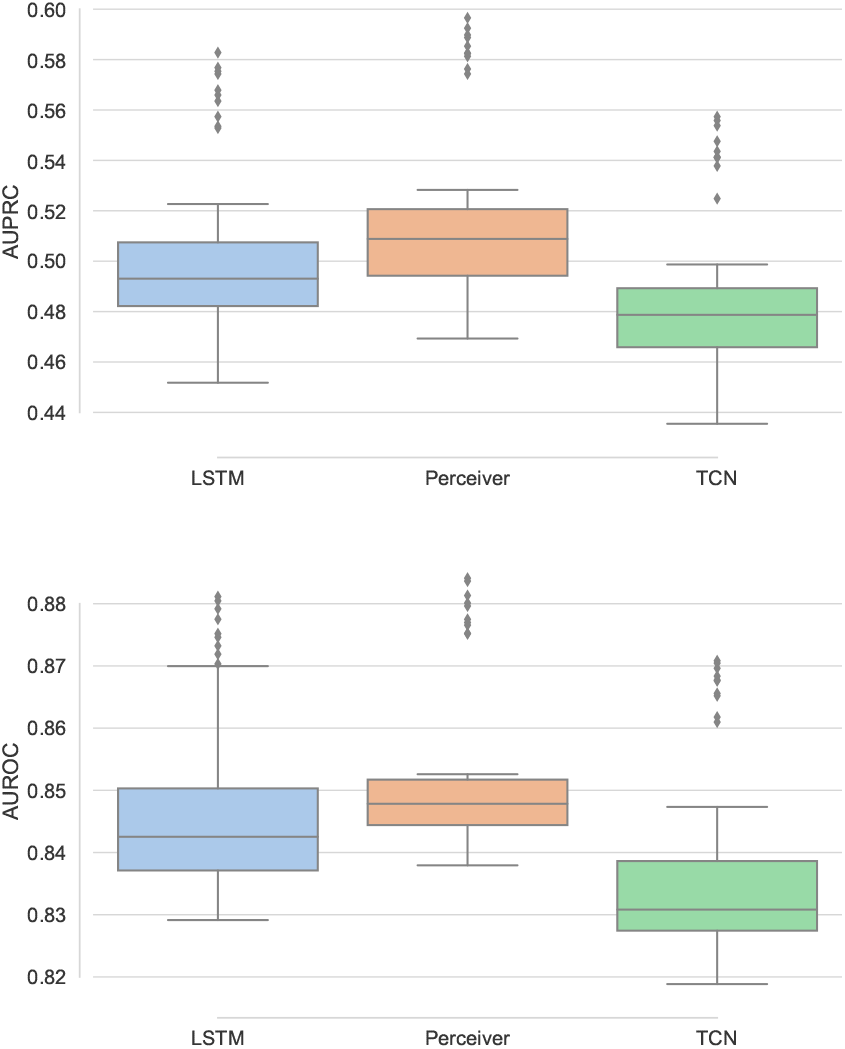
Comparative study of Perceiver against the baselines on Physionet dataset using AUPRC (upper) and AUROC (lower).

The comparative study of the Perceiver with the baselines on MIMIC dataset is presented in Fig. 4. The upper panel compares the AUPRC and as it is clear from the figure, the Perceiver slightly outperforms the baselines. Moreover, as observed earlier for the Physionet dataset, LSTM performs better than the TCN. The lower panel compares the performance using the AUROC and has results different from those observed with the Physionet as well as the AUPRC on MIMIC. All the models perform very close to each other as the maximum variation in the performance was around 0.01, and TCN performs best on average. AUROC is similar for MIMIC and Physionet datasets, although AUPRC is slightly better for Physionet than the MIMIC. The performance differences for two datasets could be attributed to the difference in number of features, data points and missingness.

Next, we present experiments to show the utility of the Perceiver over the transformer. The key idea of the Perceiver is the use of the cross-attention operation (Step 6 of Algorithm 1) to squeeze the long sequence to customised smaller learnable latents. This helps the Perceiver to manage the computational complexity as compared with the self-attention operation of the transformers, which have quadratic dependence on the input sequence and may not be able to handle the large inputs.

Fig. 3 compares the computational requirements and performance of the Perceiver and the transformer on Physionet and MIMIC datasets. The left panel presents floating-point operations per second (FLOPS), middle and right panel compare AUPRC and AUROC, for the transformer and the Perceiver with varying number of latents from one to length of the input, i.e., number of time-steps in the input (48 in our case). From the figure, we observe that by controlling the number of latents in the Perceiver, we can reduce the computations as compared with the transformer. We can reduce computations by around nine times on MIMIC and Physionet datasets, without any significant drop in the performance except AUPRC on MIMIC dataset. Transformer and the Perceiver have exactly the same architecture except for the latents introduced by the Perceiver, and because of those latents, the Perceiver takes more FLOPS for 30 to 48 latents.

**Fig. 3:**
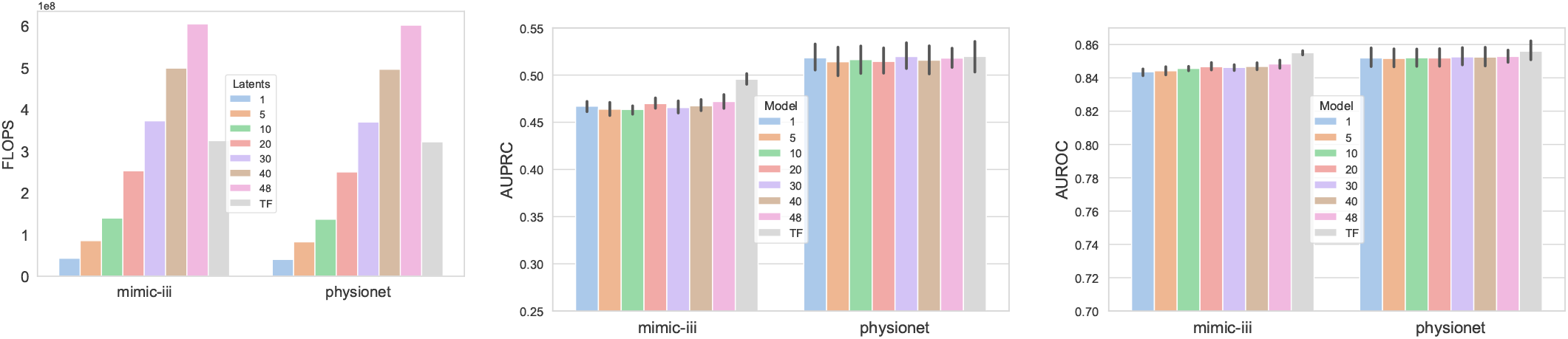
Comparative study of the Perceiver with varying number of latents and the transformer (TF) models, in terms of FLOPS (left), AUPRC (middle) and AUROC (right).

**Fig. 4:**
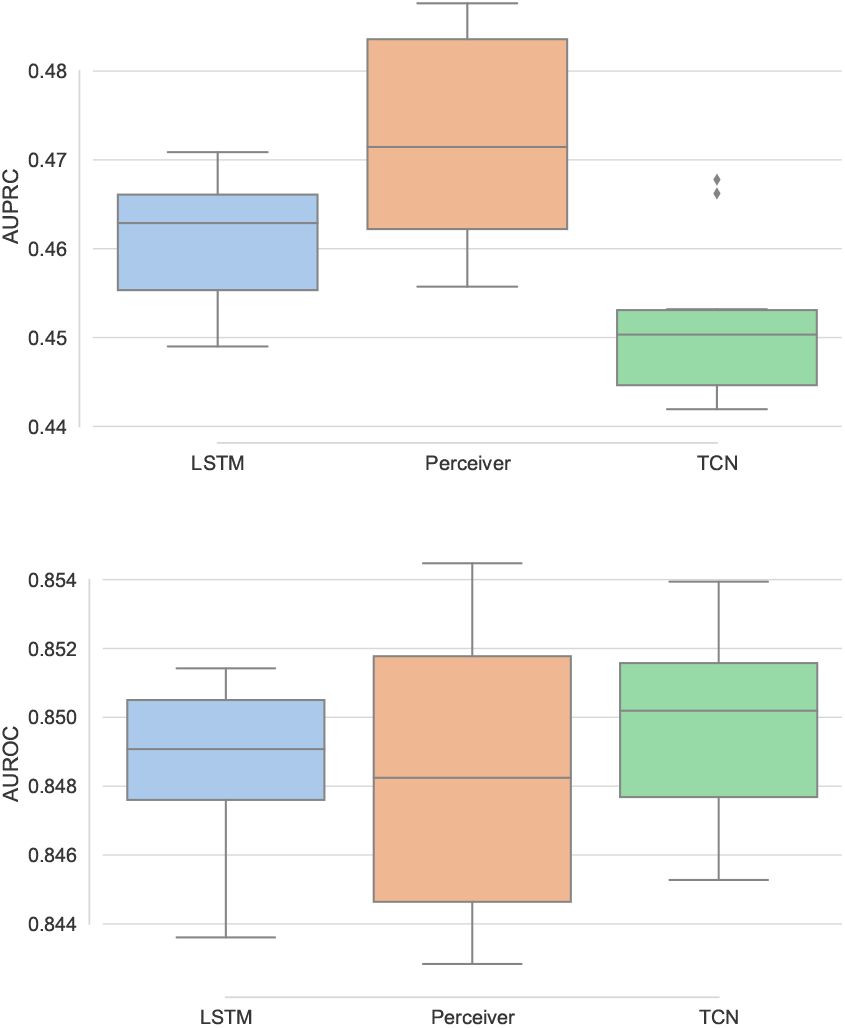
Comparative study of Perceiver against the baselines on MIMIC dataset using AUPRC (upper) and AUROC (lower).

### D. Continuous Patient State Attention

Here, we present results for the proposed continuous attention models, i.e., COPER and continuous transformer (CTransformer), to deal with the irregularity in EHR data. Continuous attention models learn the patient health dynamics from the observed irregular observations each of which representing the patient health state at a given time. Once the patient health dynamics is learned, any number of samples can be taken and at any time-step. To study the continuous models, we specifically design experiments at irregularity of 0%, 25%, 50% and 75%, and study the performance of the proposed models against the baselines, such as mTAND and LODE, as well as LSTM and Perceiver using carry forward techniques. In carry forward technique for dealing with the irregularity, we simply replace the missing steps with the previous available observation.

The comparative study of the proposed continuous patient state models against the baselines on Physionet dataset is presented in Fig. 5 using the AUPRC (upper) and AUROC (lower). From the upper panel, we find that the CTransformer is the best model and performs slightly better than the Perceiver model. Although, like LODE and mTAND, CTransformer shows large variability in the performance as compared with COPER and Perceiver models. LODE performs the worst. With the increasing irregularity in EHR, mostly the performance remains the same except a slight drop at 75%. The lower panel of the figure, compares the AUROC and have performance similar to the AUPRC. LODE and LSTM show more variability than the rest of the models. Simple baselines with carry forward, i.e., LSTM and Perceiver, also handle the irregularity quite well, however the Perceiver performs better than LSTM. This is in agreement with some of the literature [30] which shows that the carry forward works well for EHR in some settings.

**Fig. 5:**
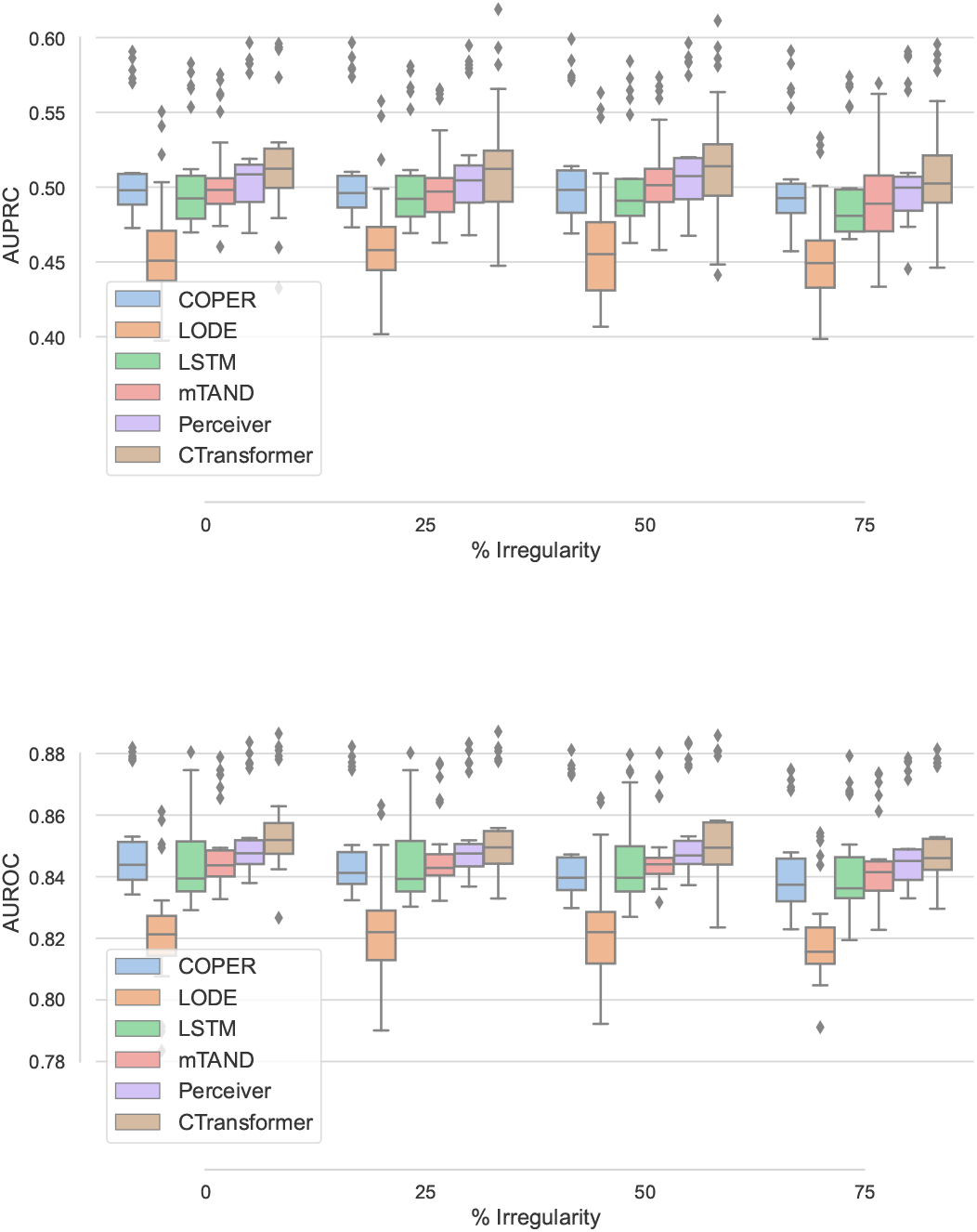
Comparative study using varying irregularity on the Physionet dataset using AUPRC (upper) and AUROC (lower).

Fig. 6 presents the performance of different models with varying degree of irregularity on the MIMIC dataset. The performance trends on MIMIC are slightly different than the Physionet. Overall, there is less variability in the results for all the models, and the variability in performance increases on the MIMIC with increasing irregularity. The proposed continuous variant of transformer, i.e., CTransformer significantly (*p <* 0.01) outperforms all the baselines and relatively shows small variability in the performance. LSTM performs the worst as it has relatively more decrease in performance as well as has more variability in the performance with increasing irregularity. Although, the Perceiver, which also uses simple carry forward mechanism like LSTM, performs with almost no drop in performance until 50% irregularity and a slight drop at 75% irregularity. Perceiver also performs better than its continuous version COPER. One potential reason for this could be over 90% missingness in the MIMIC dataset [4]. LODE performs better on the MIMIC dataset than the Physionet dataset, as it observes very small drop in performance with increasing irregularity.

**Fig. 6:**
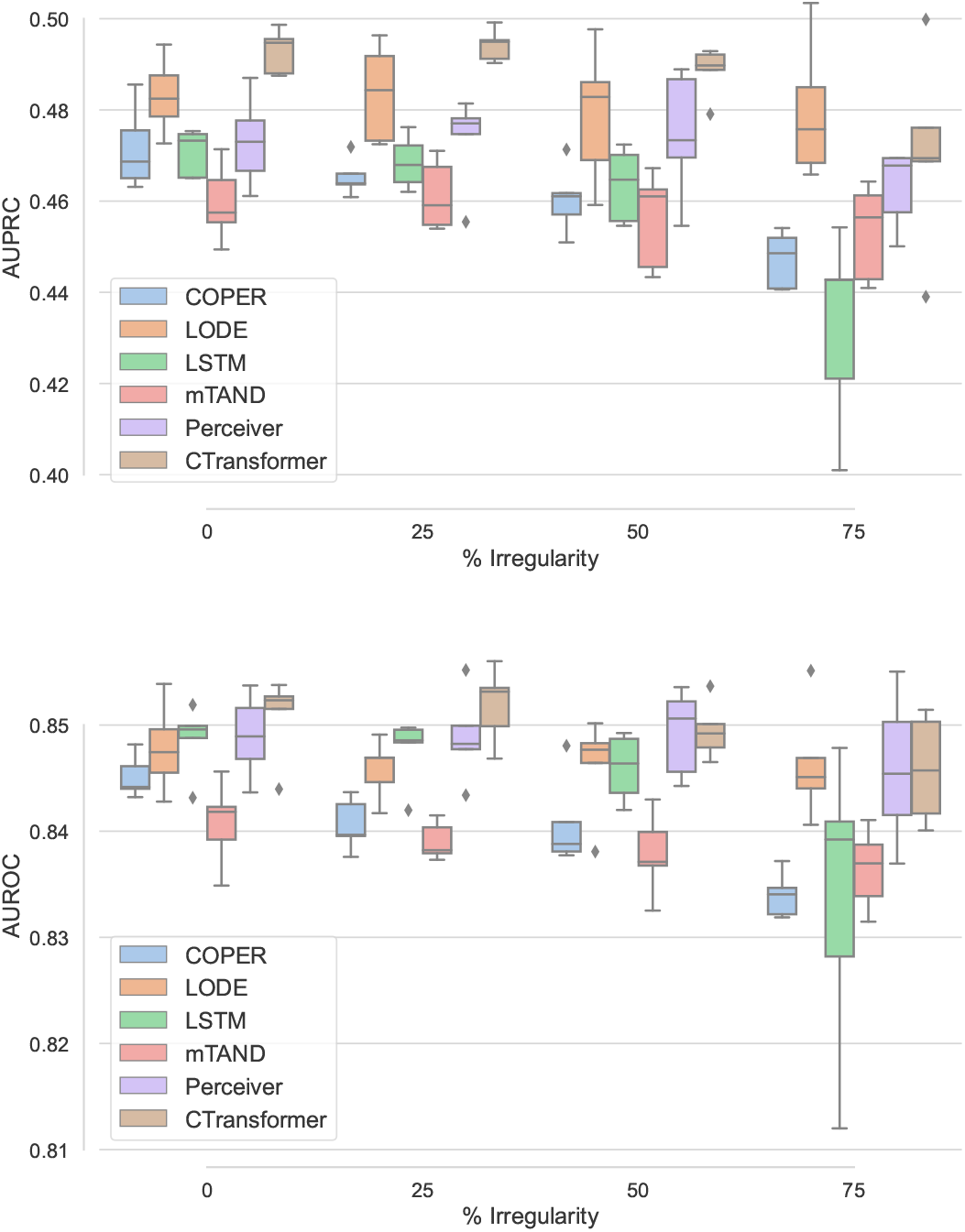
Comparative study using varying irregularity on the MIMIC dataset using AUPRC (upper) and AUROC (lower).

The techniques which utilise NODE for handling the irregularity in EHR, i.e., COPER, CTransformer and LODE are computationally extensive due to the use of the MLP in NODE. Among these, LODE uses two NODEs, one each in the encoder and decoder, and is the most computationally expensive. For the MIMIC dataset, one run of the LODE can take up to one and half day so it is not a good option for long sequences.

During the evaluation for continuous attention models, we have choice of either generating the entire time-series after learning the dynamics from the ITS or keep the observed time-steps and generate only the missing time-steps. For small irregularity and noisy data, generating the entire time-series could be helpful to reduce the effect of the noisy data.

### E. Selective Predictions and Expert Referrals

The predictive uncertainty of a machine learning model is useful in guiding the use of the model in high-stake applications, such as healthcare. The models are used for selective predictions and highly uncertain predictions of a model are referred to the expert for further examination. This will increase transparency and trust of the clinical users in the machine learning techniques and will help in the adoption of the machine learning in healthcare.

We have used Monte Carlo (MC) dropout technique [31] for calculating the model’s predictive uncertainty and is an approximation of Bayesian techniques [31] which are difficult to train. MC dropout is a simple but scalable technique and does not require training multiple models or even retraining rather trained models which use dropout for regularisation can be used for the uncertainty quantification. MC dropout requires to activate the dropout layers during the testing phase which is otherwise turned off. So each evaluation of the model with the same data point gives different prediction probabilities. We evaluated our models 25 times on each sample of the test dataset, and the mean and variance of these 25 predictions act as actual predictions and predictive uncertainty of the model.

We refer the highly uncertain cases to the experts and evaluate the model performance selectively on the remaining test dataset. Fig. 7 presents the test accuracy against the proportion of cases referred to the expert. Upper panel presents the results for the Perceiver with Physionet and lower panel presents results for the COPER with MIMIC dataset at 50% irregularity (selected randomly). Both the figures show similar behaviour and as expected, as the uncertain cases are removed, the performance of the both the models improves. Thus, the uncertainty quantification is useful and cases can be referred to clinicians as per their availability.

**Fig. 7:**
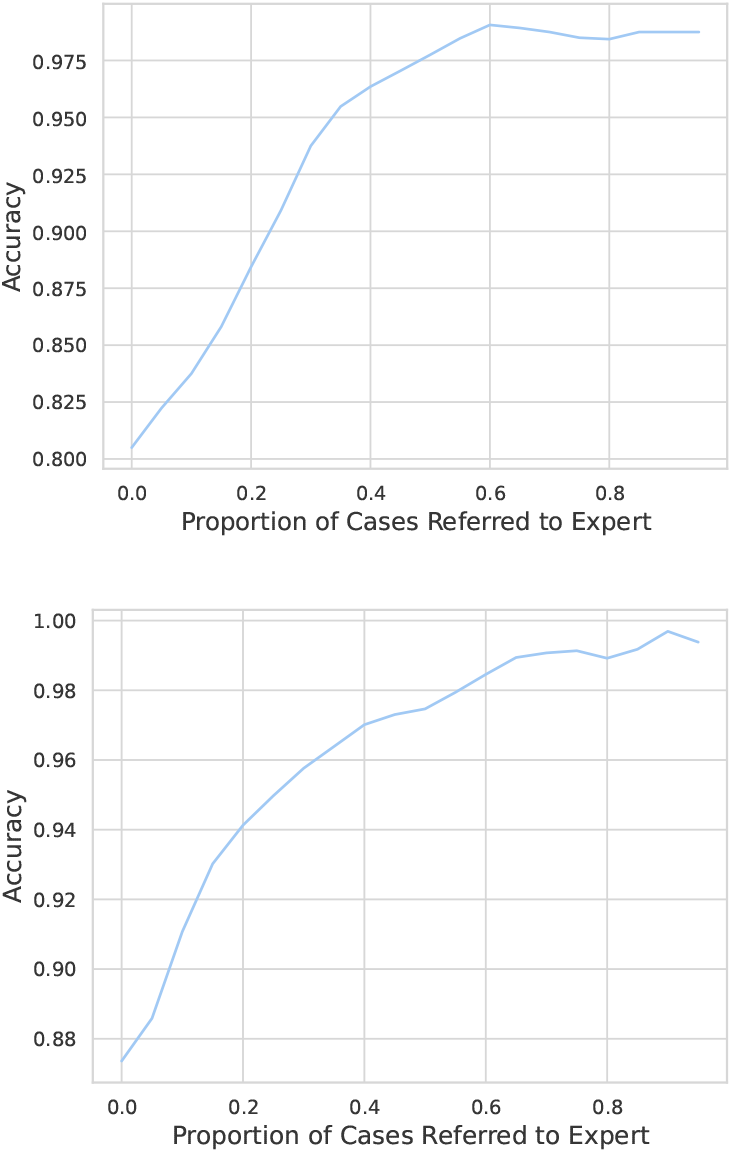
Utilisation of predictive uncertainty for selective predictions and refer the uncertain predictions to the experts: upper panel presents the test accuracy of the Perceiver on Physionet and lower panel presents the test accuracy of the COPER on MIMIC dataset, against the proportion of uncertain cases referred to the experts.

## IV. Discussions

Based on the idea of cross-attention based architectures, we proposed a computationally efficient variant of the transformer, called Perceiver, as a potential alternative for processing time-series data in EHR. The cross-attention operation helps to squeeze the long sequences of time-series to a smaller number of latents which then can be processed using self-attention operations, requiring fewer computations than directly processing the time-series with the transformer based models. Perceiver outperforms LSTM and TCN, the widely used techniques for time-series, and was able to reduce the computations by around nine times, as compared with the transformers without any significant loss of performance. We further extended the Perceiver and the transformer models to learn the patient health dynamics from the ITS. These continuous models employ neural ordinary differential equation (NODE) to model the patient health trajectory from which any number of points at any time-step can be sampled, and hence addressing the irregularity issue in EHR.

These continuous attention models can handle long sequences, completely missing time-steps, noisy observations and employ end to end learning for handling the irregularity. The experiments prove the efficacy of the proposed work on in-hospital-mortality prediction task using Physionet and MIMIC-III datasets. We also employ the uncertainty quantification for calculating the predictive uncertainty of the proposed models, which was used for selective predictions and referring the uncertain cases to the expert. This helps in improving the performance of the system, adjust the working of the models as per the time availability of the clinicians, and builds transparency and the trustworthiness of the proposed techniques for adoption in healthcare.

LSTM with carry forward technique for handling the irregularity does not perform well as it shows decrease in performance with the increasing irregularity in EHR. Moreover, the Perceiver with carry forward performs significantly better than the LSTM with carry forward for handling ITS. Overall, CTransformer outperforms all other techniques. COPER also shows competitive performance to deal with the irregularity and is computationally less expensive than the CTransformer and LODE models. Amongst all the techniques for handling ITS, LODE is the most expensive and takes up to two days to train on the MIMIC dataset. Thus, the Perceiver and the continuous patient state attention models provide computationally efficient techniques for handling ITS in EHR.

The proposed attention based models are advanced deep learning models so they share the same limitations as the other models of the same type, such as requiring more data to train, hyperparameter tuning and more computational resources than the traditional machine learning approaches. Despite this, we were able to reduce computations compared to the transformer, and the proposed models to handle ITS are computationally cheaper than state-of-the-art NODE-based models, such as LODE. To further evaluate the performance of the Perceiver and the continuous attention models, in future, we will study more tasks and datasets.

## V. Conclusions

The proposed Perceiver model provides a computationally efficient potential alternative for time-series in EHR as compared with the transformer and outperforms the commonly used baselines, such as LSTM and TCN, for in-hospital-mortality using MIMIC-III and Physionet-2012 datasets. The continuous patient state attention models, i.e., COPER and CTransformer, can handle completely missing time-steps, long sequences and provide end-to-end learning for handling irregularity in EHR. The carefully designed experiments prove the efficacy of the proposed techniques.

## Data Availability

All data produced are available online at https://physionet.org/content/challenge-2012/1.0.0 and https://mimic.mit.edu/

## Footnotes

1 https://github.com/YerevaNN/mimic3-benchmarks

2 https://github.com/locuslab/TCN

3 https://github.com/reml-lab/mTAN

4 https://github.com/YuliaRubanova/latentode

## Notes

This work was supported in part by the National Institute for Health Research (NIHR) Oxford Biomedical Research Centre (BRC) and in part by InnoHK Project Programme 3.2: Human Intelligence and AI Integration (HIAI) for the Prediction and Intervention of CVDs: Warning System at Hong Kong Centre for Cerebro-cardiovascular Health Engineering (COCHE). V. K. Chauhan is supported by a Medical Research Council (MRC) Research Grant (MR/W01761X/1).

### Competing Interest Statement

The authors have declared no competing interest.

### Funding Statement

This work was supported in part by the National Institute for Health Research (NIHR) Oxford Biomedical Research Centre (BRC) and in part by InnoHK Project Programme 3.2: Human Intelligence and AI Integration (HIAI) for the Prediction and Intervention of CVDs: Warning System at Hong Kong Centre for Cerebro-cardiovascular Health Engineering (COCHE).
V. K. Chauhan is supported by a Medical Research Council (MRC) Research Grant (MR/W01761X/1).

